# The attitudes of Greeks towards second-hand smoke and the anti-tobacco legislation

**DOI:** 10.1101/2020.11.26.20234666

**Authors:** Anna Passa, Ioannis Agtzidis, Maria Tsaousidou, Katalin Fekete Passa

## Abstract

Greece had according to Eurobarometer 2017 the highest rates of secondhand smoke exposure in the European Union (87%). The main aim of this study was to understand the reasons for the non-enforcement of the anti-tobacco legislation. To do this, we created two different questionnaires, one for smokers and one for non-smokers, and we collected epidemiological data, data about the attitudes of Greeks towards smoking, second-hand smoke, and the smoke-free legislation, as well as data about some relevant behavioural patterns. In total 597 non-smoker questionnaires and 366 smoker questionnaires were collected, with the mean age of the participants being 40 years old. The majority of people claimed that smoke disturbs them and, interestingly, smokers responded that they consider disturbing others with their smoke, and especially when children are present. Additionally, smokers said they would slightly reduce going out if the smoke-free legislation was strictly enforced, while non-smokers would respectively slightly increase going out. Based on this observation and given the higher proportion of non-smokers than smokers, we can assume that there will be no negative impact to the eating and drinking establishments, as many people were speculating, but to the opposite, there might even be a small benefit from the implementation of smoke-free measures. Since 2019 the relevant legislation has been enforced, but due to the COVID-19 pandemic, the long-term implementation and the true outcomes of the relevant legislation are more difficult to study.

## Introduction

Smoking and exposure to second-hand smoke (SHS) are some of the largest avoidable health risk factors and they contribute substantially to the overall percentage of premature deaths. Around 50% of smokers die prematurely (1), resulting in a decrease of an average smoker’s lifespan by 14 years. Because of the severity of many common health problems that relate to long-term smoke exposure, which among others include lung cancer, lower respiratory tract infections, otitis media, and ischemic heart disease, the European Union and its member states proceeded with a prohibition of smoking in indoor and public spaces. On 30^th^ November 2009, the European Commission with its 2009/C 296/02 recommendation (2) strengthened the smoke-free legislation within public spaces, as well as EU cooperation on tobacco control. The implementation of the new legislation in many EU member states dropped the overall smoke exposure rate of the EU citizens in bars and pubs from 46% in 2009 to 28% in 2012, and in restaurants from 31% to 14% accordingly (3)(4). A newer survey of the Eurobarometer in 2017 showed that the exposure rates had dropped further to 20% in bars and pubs, and to 9% in restaurants (5). The new policies also resulted in a reduction of smokers in the EU between 2006 and 2017 and especially in the period of time before 2014. In total there has been a six percentage point decline in the proportion of those who smoke (6) and their percentage in 2017 stood at 26%. Nevertheless, the rates of smokers and of secondhand smokers (people that do not smoke themselves but are exposed to cigarette smoke) have barely changed and are still high in some EU countries (7) (5).

More specifically, in Greece according to the Special Eurobarometer 458 of 2017, 37% of the overall population were smokers, 87% of the citizens were exposed to SHS in drinking establishments such as bars, and 78% of the citizens were exposed to SHS in eating establishments such as restaurants. These numbers across the EU were 20% and 9% respectively. This vast difference is the result of the non-enforcement of the relevant legislation. This non-enforcement could be partly attributed to the owners of eating and drinking establishments as well as part of the Greek population, both of whom have complained about the anti-tobacco legislation. The association of cafe, bar, restaurant, and club owners have also protested against this legislation, due to their fear of losing smoking customers (8)(9)(10)(11)(12)(13)(14)(15)(16). Nevertheless, the main aetiology behind the non-enforcement remains uncertain.

Greece has a long history of smoke-free legislation with the first one having been implemented in 1856. Several other non-smoking legislation followed during the twentieth century, as well as in the past two decades, with most of them being poorly enforced for many years (A detailed list of the smoke-free legislation in Greece can be found in Table 1). The last legislation of 2019 seems to have a long-term effect for the first time and potentially the high exposure to secondhand-smoke might have dropped during the past year. The main aim of this study is the investigation of the attitudes of Greek people towards smoking in indoor spaces in general and towards the smoke-free legislation, in order to better understand the reasons behind the non-enforcement of anti-tobacco legislation until 2019. In this article, we provide data about the personal smoking habits of greeks, their opinion about smoke and secondhand smoke, and their willingness to visit areas where smoking is prohibited. Then we present the statistically significant results from the correlation analysis between some of the research points of the questionnaires and the question regarding the possible change of the going-out-habits of Greeks if the smoke-free legislation was strictly obeyed. All the gathered data together with the questionnaire forms are provided in https://gin.g-node.org/ioannis.agtzidis/smoking_survey.

**Table 1.** Smoke-prohibiting legislations

| Year | Legislation goal |
| --- | --- |
| 1856 (17) | Prohibition of smoke in public offices and shops |
| 1952 (18) | Prohibition of smoke in public transport |
| 1956 (19) | Prohibition of smoke in theatres and cinemas |
| 1979 (20) | Prohibition of smoke in healthcare-institutions and private clinics |
| 1980 (21) | Prohibition of smoke in public indoor spaces |
| 1990 (22) | Prohibition of smoke on domestic flights |
| 1993 (23) | Prohibition of smoke in healthcare environments |
| 1995 (24) | Prohibition of smoke on passenger ships |
| 2002 (25) | Prohibition of smoke in public spaces, public transport, and healthcare environments |
| 2005 (26) | Acceptance-implication of the WHO Framework Convention on Tobacco Control (United Nations 13.06.2003)/ |
| 2007 (27) | Prohibition of smoke in indoor playgrounds |
| 2008 (28) | Protection of children, all indoor spaces, eating and drinking establishments, |
| 2010(29)(30) | Prohibition of smoke in public spaces and banning of tobacco product advertisement |
| 2011 (31)(32) | Law aiming to enforce the smoke prohibition law of 2010 (29) and directions about the law enforcement |
| 2013 (33) | Law aiming to enforce the smoke prohibition law of 2010 (29) |
| 2014 (34)(35) | A reminder of the reinforcement of the smoke prohibition laws from 2000 until 2013 |
| 2016 (36) | Adjustment of the domestic legislation according to the 2014/40/EU directive regarding smoke-relevant advertisements |
| 2017 (37) | A reminder of the enforcement of the 2016 legislation (36) |
| 2018 (38)(39) | Enforcement of the smoke-free legislation from 2002 until 2017, a reminder of the previous 2018 smoke-free legislation enforcement (38) |
| 2019<br>(40)(41)(42)(43) | Enforcement of the smoking prohibiting legislation, regulations regarding the enforcement, the controls and the ban of the smoke-free legislation<br>**(legislation with a long term effect)** |

\*legislation whose main aim was the prohibition of smoking and not every legislation containing smoke-free provisions (ex. fire hazard)

**Table 2.**
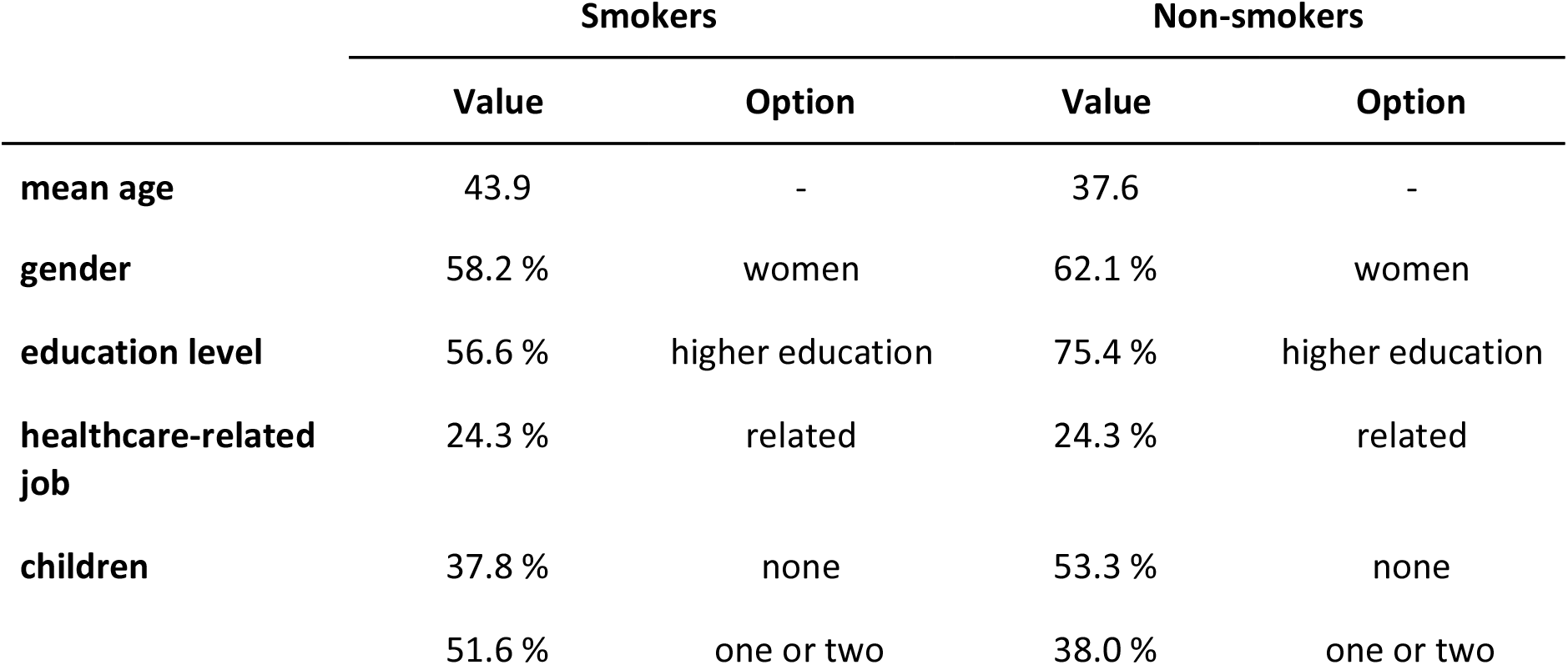
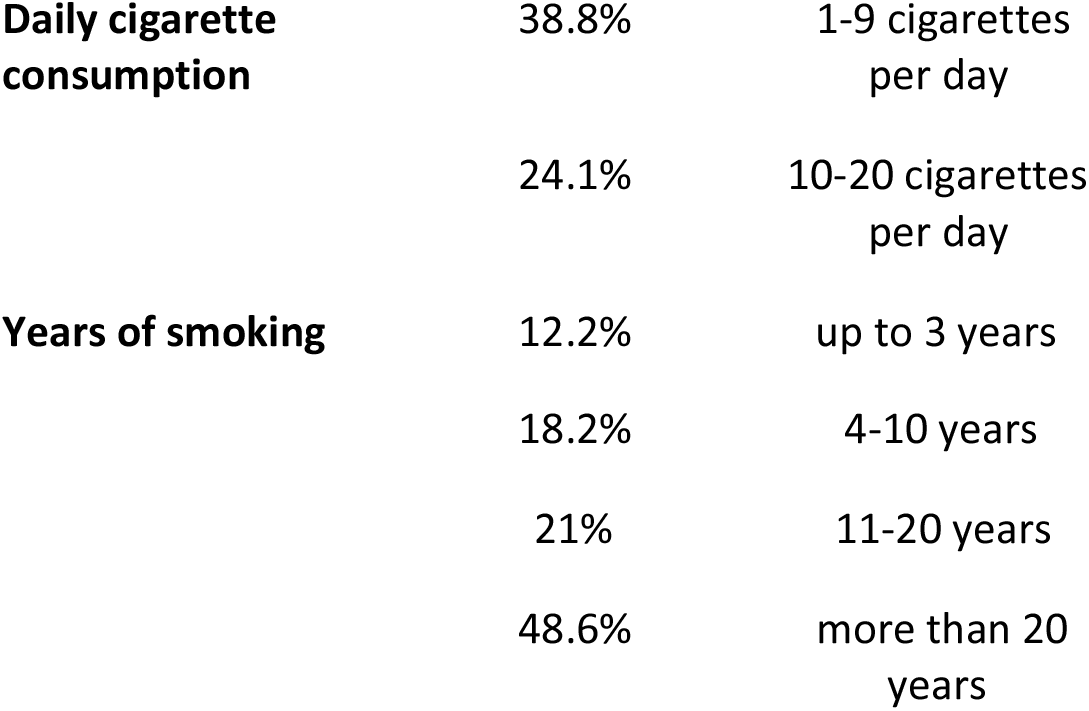
Epidemiological data

|  | Smokers |  | Non-smokers |  |
| --- | --- | --- | --- | --- |
|  | Value | Option | Value | Option |
| **mean age** | 43.9 | - | 37.6 | - |
| **gender** | 58.2 % | women | 62.1 % | women |
| **education level** | 56.6 % | higher education | 75.4 % | higher education |
| **healthcare-related job** | 24.3 % | related | 24.3 % | related |
| **children** | 37.8 % | none | 53.3 % | none |
|  | 51.6 % | one or two | 38.0 % | one or two |

|  |  |  |
| --- | --- | --- |
| **Daily cigarette consumption** | 38.8% | 1-9 cigarettes per day |
|  | 24.1% | 10-20 cigarettes per day |
| **Years of smoking** | 12.2% | up to 3 years |
|  | 18.2% | 4-10 years |
|  | 21% | 11-20 years |
|  | 48.6% | more than 20 years |

## Methods

### Data collection procedure

The present study is a cross-sectional study conducted in the wider area of Thessaloniki, in Greece. The target population included adults (≥18 years old) of all genders. Both smokers and non-smokers were picked-up randomly and participated in this survey by filling out an anonymous questionnaire. The questionnaires were distributed in classrooms of the Aristotle University of Thessaloniki, in public service waiting rooms (ex. Hospitals), and in cafés during a 12-month period (March 2018 to February 2019).

The questionnaires were created and automatically processed with the SDAPS tool (version 1.9), which eliminated the manual importation of the filled questionnaires and resulted in a reduced error rate. As a further quality assurance step, we inspected all the automatically detected questionnaire responses and corrected the false detections.

### Questionnaire structure

For this study, we created separate questionnaires for smokers and non-smokers, each consisting of two parts. The first part contained questions about demographics, and the second part contained questions regarding the participant’s opinion and attitude towards smoking, the exposure to second-hand smoke, and the prohibition of smoking in drinking and eating establishments.

The epidemiological data for both smokers and non-smokers included their age (18-25, 26-35, 36-55, 56-75, other), gender (male, female, other), educational status (primary, secondary, higher), employment in a healthcare-related occupation (yes, no) and the number of children (0, 1, 2, 3, 4+). Smokers were also asked about their daily cigarette consumption (0-9, 10-20, 21+) and the number of years they have been smoking (0-3, 4-10, 11-20,20+).

The second part of the questionnaire was the main part that was investigating the perception of the participant towards smoking indoors, second-hand smoke, and the smoke-free legislation. This section investigated mainly qualitative variables and had some common and some different questions between smokers and non-smokers. Questions regarding the probability of the participant visiting a non-smoking eating or drinking establishment, tax-sufficiency on tobacco products, smoking at home, health issues caused or burdened by smoke, lack of control of the smoke-free law, annoyance by smoke, and opinion about the harmfulness of smoke were asked to both smokers and non-smokers. Additionally, smokers were asked about the importance that harming or disturbing other adults and children has for them, as well as about their tobacco product consumption in front of their children or during pregnancy. The opinion of smokers about e-cigarettes was also investigated.

### Statistical analysis

The Statistical analysis was performed with the Statistical Package for the Social Sciences (IBM SPSS Statistics 25). The questionnaire consisted of two types of questions (namely, categorical and ordinal). For categorical data, we provide the percentages for each category, and for ordinal data, we also provide the mean and standard deviation among the available choices. Pearson’s *χ*^2^ test was used to probe the association between two questions. Results were considered statistically significant when the p-value was lower than 0.05. To check for the kind of association between the two groups (smokers vs. non-smokers), we compared the frequencies by applying a z-test on the values of the previously computed Pearson’s *χ*^2^ tests.

## Results

The results are presented for both smokers and nonsmokers either as percentages, median answer values, or mean values with standard deviation. First, we analyze the descriptive statistics, and then we present the significantly associated questions with their *χ*^2^ statistics, computed with Pearson’s *χ*^2^ test. For each question, the corresponding question number in the questionnaire is also presented in a parenthesis, where “S.” stands for smokers, “NS.” for non-smokers, and 2 or 3 numbers for the question number. The first number stands for the questionnaire part and the second (and third) for the question number (S. 2_7_3 = smokers, part 2, question 7_2, NS. 2_3 = non-smokers, part 2, question 3).

### Epidemiological data

Overall, we collected 366 questionnaires for smokers and 597 questionnaires for non-smokers. The mean age of smokers was 43.9 years (SD = 14.5 years) (S. 1_1) and the mean age of non-smokers was 37.6 years (SD = 16 years) (NS. 1_1). 58.2% of the smokers and 62.1% of the non-smokers were women (NS. 1_2)(S. 1_2) and among smokers, 56.6% had higher education (S. 1_4), whereas the same percentage for non-smokers was 75.4% (NS.1_4). The percentage of the participants working or studying in the domain of health care was the same for both smokers and non-smokers: 24.3% (S. 1_3, NS. 1_3). Most of the smokers (51.6%) had one or two children and 37.8% of them had no children (S. 1_5), while the majority of non-smokers (53.3%) had no children and 38% of them had one or two children (NS. 1_5). Smokers were additionally asked about the number of cigarettes they smoke per day, and 38.8% of them smoke 1-9 cigarettes per day, while 42.1% of them smoke 10-20 cigarettes per day (S. 1_6). Regarding years of smoking, 48.6% of smokers claim they have been smoking for more than 20 years, 21% of them for 11 to 20 years, 18.2% of them for 4 to 10 years, and 12.2% of them for up to 3 years (S. 1_7).

### Attitudes towards the smoke-free legislation

The inspection of how the smoking prohibition could affect the willingness of the participant to go out was made with rating questions whose possible answers ranged from 1 to 5 (“not at all” to “totally”). Smokers seemed to be neutral (median: 3, mean: 3.00, SD: 1.56) when asked how likely it would be for them to visit a non-smoking eating or drinking establishment (S. 2_1), they would slightly decrease going out (median: 1, mean: 2.21, SD:1.45) if the smoke-free law was everywhere strictly obeyed (S. 2_4), and would slightly decrease going out (median: 1, mean:2.09, SD: 1.41) in a foreign country where the smoke-free law is strictly controlled (S. 2_5). Non-smokers would on the other hand moderately increase going out (median: 3, mean: 2.67, SD: 1.48) if the smoking prohibition was strictly obeyed (NS. 2_4), and would moderately increase going out (median: 3, mean: 2.76, SD:1.51) when visiting a foreign country where the smoke-free law is strictly controlled (NS. 2_5). The mean values of the participants’ choices are presented in Figure 1.

**Figure 1.**
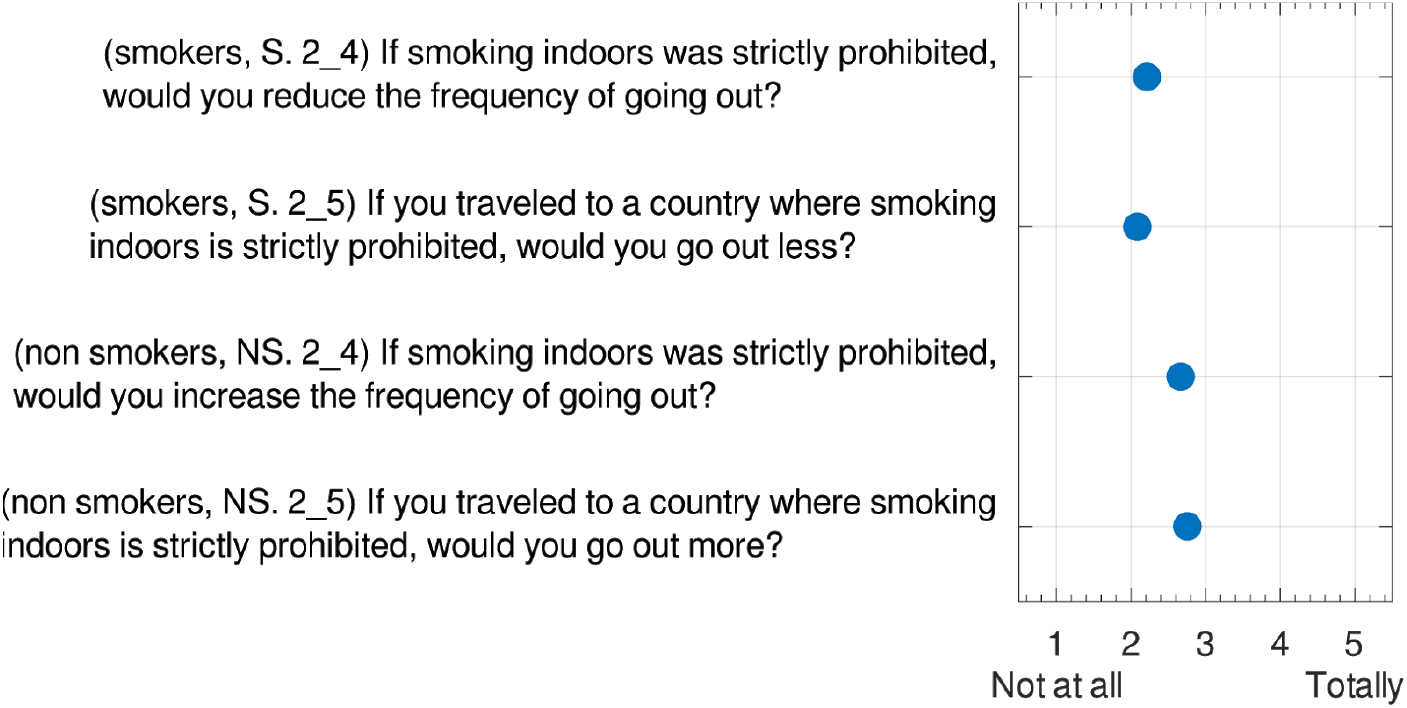
“Going out” related to smoke-free policies

### Attitudes towards smoking and second-hand-smoke

Weekly hours that non-smokers spend in indoor spaces where the smoke-prohibition is not obeyed (NS. 2_2), showed that 49.2% of them spend weekly 0-4 hours in closed spaces where smoke-free law is not obeyed, 33.4% of them spend 5-10 hours and 9.4% of them spend 11-20 hours with the rest spending weekly more than 20 hours. Regarding the complaints that non-smokers might have made about the smoke during the last year (NS. 2_3), 35.9% of them claimed they have not made any, while most of them (41.9%) have complained 1-5 times. Smokers were respectively asked whether they have received complaints about them smoking during the last year, and 57.6% of them claimed that they have not received any complaints while 28.7% of them have received 1 to 5 complaints (S. 2_3). Smokers were additionally asked about the importance that disturbing others with their smoke has for them(S. 2_2), and they claimed they take a lot under consideration that they might disturb people around them (median:4, mean:3.95, SD: 1.28). Smokers also claimed to take a lot under consideration (median: 5, mean: 4.47, SD: 1.09) that they might harm children in the same closed area (S. 2_6).

The last part of the questionnaire consisted of closed-ended type questions, to which the participants could choose between “yes” or “no”. The results showed that the majority of the participants, 95% of the smokers and 98.6% of the non-smokers, believe that smoking (for smokers) and second-hand smoke (for non-smokers) is harmful (S. 2_7_9, NS. 2_1_2). The majority of the participants, 57.2% of smokers and 87.6% of non-smokers are also being disturbed by smoke (S. 2_7_1, NS. 2_1_1). Some have also already had an illness caused or burdened by smoke. This percentage was 19.3% for smokers and a little lower (13.8%) for those who do not smoke (S. 2_7_12, NS. 2_6_6). Questions regarding the smoking-allowance at home showed that 48.3% of the smokers smoke at home (S. 2_7_4), and 60.7% of them let their guests smoke at home (S. 2_7_7). Similar questions for non-smokers showed that 34.1% of them have somebody that smokes at home (NS. 2_6_3), and 37.9% of them let guests smoke in their home (NS. 2_6_4). A small percentage of non-smokers (4.6%) said they smoke occasionally when they go out (NS. 2_6_5). Smokers were also asked about their smoking habits in front of their children, and 55.5% of them claimed that they do smoke in front of their children (S. 2_7_5). Women who smoke were asked about smoking during pregnancy too, and 23% of them said they did smoke during pregnancy (S. 2_7_6). Questions regarding the hypothetical perception of the situation in Greece, as far as smoking indoors, if the participants were tourists, showed that 70.8% of the smokers would not find the situation positive (S. 2_7_3), while 90.6% of the non-smokers would find the situation negative (NS. 2_6_2). Taxes on tobacco products were considered insufficient for the expenses of the health care system caused by smoking by 72.2% of the smokers (S. 2_7_2) and 79.4% of the non-smokers (NS. 2_6_1). At the end of the closed-ended type questions part, smokers were asked about their perception of electronic cigarettes and 79.4% of them appeared to believe that e-cigarettes have a negative impact on health (S. 2_7_10), and only 38.2% of them think that electronic cigarettes harm less than tobacco products (S. 2_7_11). The mean value of the responses of the participants to the closed-ended type questions is presented in Figures 2 and 3.

**Figure 2.**
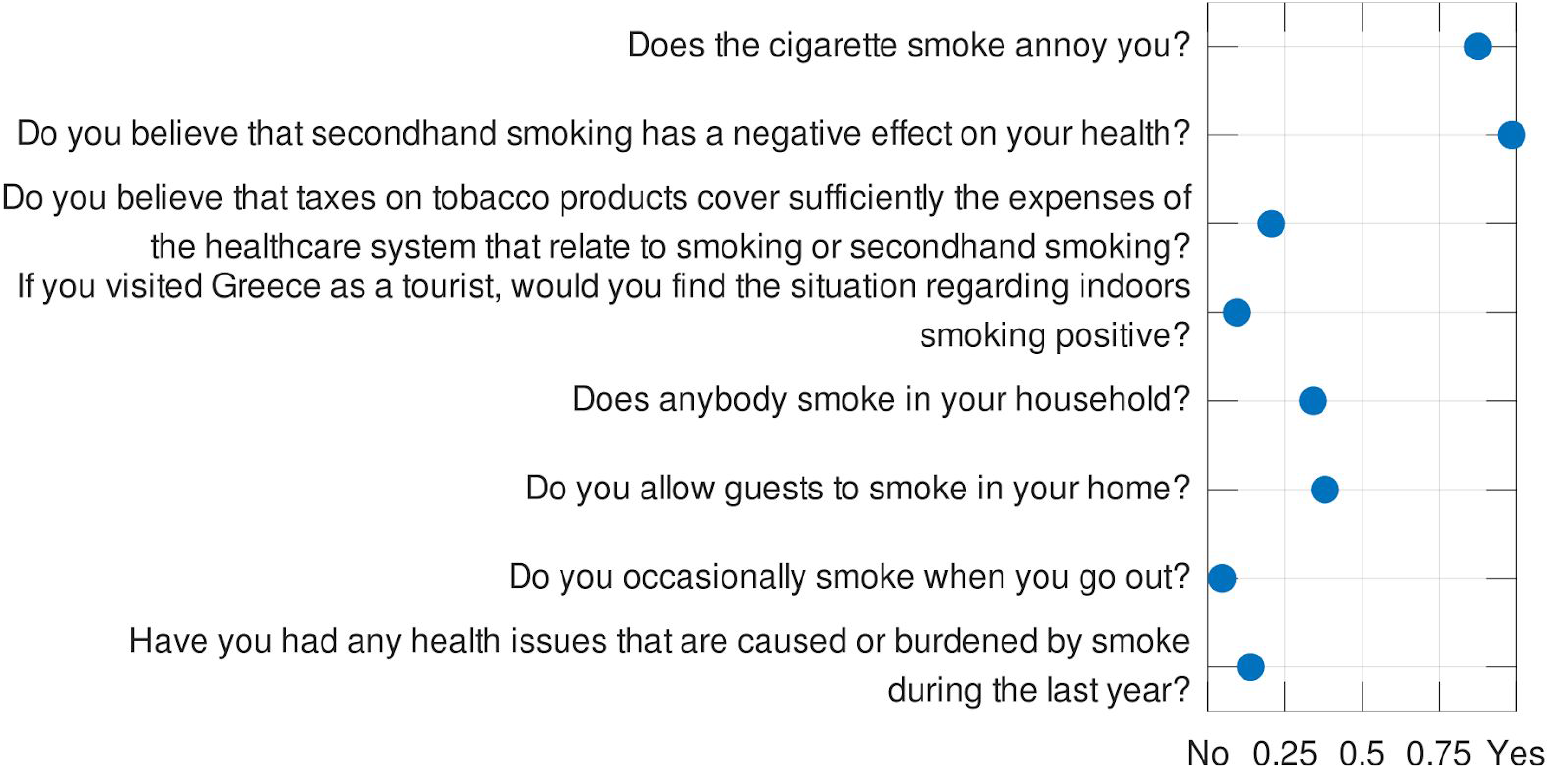
Non-smokers - Closed-ended type questions: mean value of the answers

**Figure 3.**
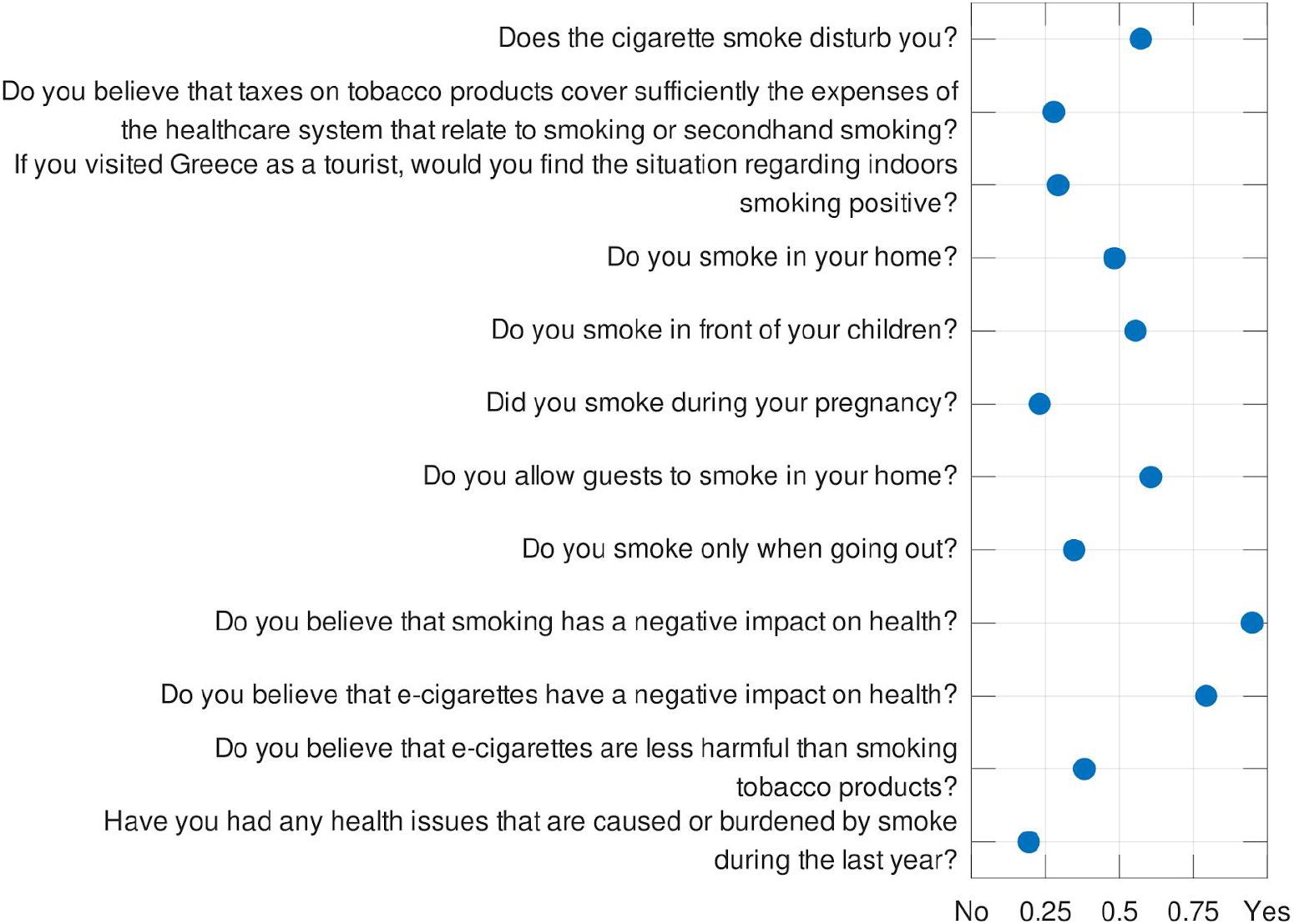
Smokers - Closed-ended type questions: mean value of the answers

### Association between the willingness to go out and other questionnaire parameters

As far as association is concerned, Pearson’s *χ*^2^ test was conducted to probe for the association between the questions regarding the possible changes in the participant’s going-out-habits (S. 2_4, NS. 2_4), and all the other questions of the questionnaire. As next, we present the significantly associated questions with the probable increase (for non-smokers) or decrease (for smokers) in going out, if the smoke-prohibition was everywhere strictly obeyed.

Smokers were asked if they would decrease going out if the smoke-free legislation was strictly obeyed, and Pearson’s *χ*^2^ test showed a significant association between this decrease and the age class of the participants (*χ*^2^ = 27.765). Youths, aged between 18 and 25 years old, had an average of 1.72 on the rating scale, while people aged between 26-35 years old and the class of middle-aged people (36-55 years old) had an average of 2.2 and 2.45, respectively. Older people, aged between 56 and 75 years old, were less likely to reduce going out (mean value = 2.08) when compared to their previous generation. This means that the youngest and the oldest participants would be less affected in going out in comparison to those in the middle.

The number of cigarettes a smoker smokes per day is also statistically significantly associated with the decrease in going out (*χ*^2^ = 33.693, p < 0.05). The association is positive, meaning that the more cigarettes one smokes the more likely he/she is in decreasing going-out. Additionally, tobacco product taxation sufficiency is statistically associated with decreasing going-out (*χ*^2^ = 9.886, p < 0.05). This means that smokers who find the tobacco products taxation sufficient for the expenses that occur at the health care system from the tobacco product consumption have a bigger tendency to decrease going out with the ban of smoking indoors. Additionally, women who smoked during pregnancy are going to reduce going out more, in comparison to the ones who did not smoke in the same period (x^2^ = 9.749, p < 0.05).

Respectively, non-smokers were asked if they would increase going-out in case smoke-free legislation was strictly obeyed. Pearson’s *χ*^2^ test (*χ*^2^ = 58.707, p < 0.05) showed a statistically significant association between the frequency of going out more after strict smoking prohibition and age. Youths between 18 and 25 years old had an average of 2.18, the age class 26-35 had a mean value of 2.94, people with 36-55 years of age had a mean value of 3.09, and people between 56 and 75 years old had a mean value of 2.84 on the rating scale. This means that youths (18-25 years old) would not go out more while the rest of the non-smokers seem as they would moderately increase going-out.

For non-smokers, the number of children was also associated with increasing going out if the smoke-free legislation was strictly obeyed (*χ*^2^ = 54.246, p < 0.05). Non-smokers with 1 or more children seem to moderately increase going out while the ones without kids would only increase going out a little. The same variables were not significantly associated with smokers. The subjective annoyance of a non-smoker by smoke had also a statistically significant association with him/her increasing going out if smoke prohibition was strictly obeyed (*χ*^2^ = 52.363, p < 0.05). This means that people who are bothered by smoke would go out more after law obeyance. Non-smokers who declared that they do not let their guests smoke in their homes are more likely to go out more in smoke-free circumstances (*χ*^2^ = 49.357, p < 0.05). The fact of whether somebody smokes at home or not was also proved significantly associated with the possibility of non-smokers going out more (*χ*^2^ = 12,084, p < 0.05). More specifically, non-smokers who did not have somebody smoking at home were more likely to increase going out with the strict restriction of smoking indoors. Some non-smoker participants who may smoke when they go out were also less likely to increase going out compared to the non-smokers who never smoke (*χ*^2^ = 13.699, p < 0.05). Additionally, the number of complaints a non-smoker made is highly and statistically associated with the possibility of him/her going out more (x^2^ = 73.544, p < 0.05), which means that the more complaints one made the more likely he/she is to go out more after smoking restriction law applies.

Sex, working in the healthcare system and education level are variables that did not have an association with increased or decreased going out neither for smokers nor for non-smokers. Nevertheless, there was a significant association between education level and the probability of smoking. Participants with primary or secondary education level had a higher chance of being smokers, contrary to participants with higher education, who were less likely to be smokers. (*χ*^2^ = 38.921, p < 0.001).

## Discussion and conclusion

The high rates of smokers and smoke exposure in Greece have provoked the scientific community to widely research the smoking prevalence, exposure rates to second-hand-smoke, as well as the epidemiologic and socioeconomic characteristics of smokers in Greece (44)(45)(46)(47)(47,48)(49)(50). However, only a few studies researching the attitudes of Greek people towards the smoke, second-hand smoke, and smoke-free legislation have been conducted. The Greek institute of public health and the American College of Greece, in cooperation with the polling company KAPA Research, has conducted in 2017, and in 2019 some of the few studies regarding the attitudes of Greek people towards the non-enforcement of the anti-tobacco legislation. The study in 2017 showed that 88.1% of the Greek population was against smoking and 83.8% considered exposure to secondhand smoke as a cultural degradation, while 96.6% had been exposed to secondhand smoke and 93% believed that the state has not taken sufficient measures to control smoking in indoor spaces (51). Another study of 2019 showed an overall secondhand smoke exposure of 94.6% and a high percentage of the Greek population (87.5%) wanting the smoke-free legislation to be enforced (52).

Nevertheless, the behavioural patterns of Greek people as far as smoking is concerned, as well as their behavioural changes in relation to smoking-prohibition, are some factors that had not been properly investigated until now. In this study, we investigate the attitudes of Greeks towards the smoke-free legislation and second-hand-smoke with respect to the potential changes of their going-out habits and to the above-mentioned behavioural patterns. These parameters do influence the imposition of the anti-tobacco measures and understanding them could substantially contribute to the identification of the reasons, real or fictional, that resulted in the inefficient application of the anti-tobacco policies and thus improve future policy decisions.

With our questionnaires, we tried to cover all the previously mentioned topics but there were some weaknesses in the process. We specialized questions on the same topic between smokers and non-smokers, which sounded like a good idea initially but ultimately made the analysis and direct comparison between smokers and non-smokers more difficult. One such example is the question regarding the going-out habits if smoking was strictly forbidden. Also performing this study required a long period of time for the data gathering (since February 2018) and the data analysis. During this period the law prohibiting smoking in indoor spaces in Greece has been implemented (autumn 2019) to an extent that makes the results of our study important only at a theoretical level and in some places where the debate about smoke-free spaces still exists.

But most importantly our data help to debunk one of the main argument against the uniform enforcement of the anti-tobacco policies: the fear of the owners of drinking and eating establishments regarding the potential loss of customers (8) (9) (10) (11) (12) (13) (14) (15) (16). These speculations along with the fear of politicians becoming unpopular (53)(54) had resulted in a long history of ever stringent smoking legislation not being enforced (see Table 1). Given our results, we can understand that the enforcement of the anti-tobacco policies wouldn’t have the negative effects that many people were speculating. When taking into consideration the fact that non-smokers constitute a considerably bigger portion of the population (prevalence of smokers in Greece according to Special Eurobarometer 2017: 37% (5)), we can assume that the number of clients in drinking and eating establishments would follow a similar pattern. Based on this observation and on our results that show a slight increase in going-out rates for non-smokers versus a slight decrease in going-out rates for smokers (see Figure 1) we can expect no negative effect but to the contrary even a slight increase in clientele with the strict and universal prohibition of smoking in indoor spaces. Given the fact that other studies report an even lower percentage of smokers (approx. 27%) (51) (52) than the Eurobarometer of 2017, this could point to an even bigger change in the opposite direction of the concerns of the owners of eating and drinking establishments. Now, since the smoke prohibition has been implemented it would be of interest to investigate the actual outcomes, as far as the going-out habits and the satisfaction of people are concerned. Nevertheless, an investigation like this would be very difficult in the current situation because the coronavirus pandemic has vastly changed our going-out habits and its effects are probably going to be long term.

## Summary

We gathered data about the attitudes of Greeks towards smoking, secondhand-smoke and the smoke-free legislation from the Thessaloniki metropolitan area, from March 2018 to February 2019. We performed an initial analysis of all our data and then we looked for a correlation between each parameter of our questionnaire and the data regarding the going out habits of respondents. This analysis showed no negative effect on the going-out habits of Greeks if the legislation were enforced. All the gathered data together with the questionnaire forms are publicly available at https://gin.g-node.org/ioannis.agtzidis/smoking_survey.

## Data Availability

All the gathered data together with the questionnaire forms are publicly available.

https://gin.g-node.org/ioannis.agtzidis/smoking_survey

